# Metabolic syndrome among type 2 diabetic patients in Sub-Saharan African countries: a systematic review and meta-analysis

**DOI:** 10.1101/2020.05.14.20101410

**Authors:** Wondimeneh shibabaw shiferaw, Tadesse Yirga Akalu, Mihretie Gedefaw, Yared Asmare Aynalem, Denis Anthony, Ayelign Mengesha, Worku Misganaw, Henok Mulugeta, Getenet Dessie

## Abstract

**Background:** Metabolic syndrome is one of the serious public health problems among type 2 diabetic patients. Despite a number of studies have been conducted, there is no overall estimation on the prevalence of metabolic syndrome among type 2 diabetic patients in Sub-Saharan African countries. Therefore, this study aimed to estimate the pooled prevalence of metabolic syndrome in patients with type 2 diabetes mellitus in Sub –Saharan African countries.

**Methods:** PubMed, Web of Science, African Journals Online, Google Scholar, Scopus, and Wiley Online Library databases from inception to April 27, 2020 were searched to identify relevant studies. The I^2^ statistic was used to check heterogeneity across the included studies. DerSimonian and Laird random-effects model was applied to estimate pooled effect size, and 95% confidence interval across studies. A funnel plot and Egger’s regression test were used to determine the presence of publication bias. Sensitivity analysis was deployed to determine the effect of a single study on the overall estimation. All statistical analyses were done using STATA™ Version 14 software.

**Result:** In this meta-analysis, a total of 23 studies with 6,482 study participants were included. The estimated prevalence of metabolic syndrome in Sub-Saharan African countries was 59.62% (95% CI: 52.20, 67.03). Based on the subgroup analysis, the highest prevalence of metabolic syndrome (61.14%, 95% CI: 51.74, 70.53) was reported in Ethiopia. . Additionally, the highest prevalence of metabolic syndrome was reported across studies using the diagnostic criteria of National Cholesterol Education Program Adult Treatment Panel III 64.8% (95% CI: 54.74, 74.86), followed by International Diabetic Federation (57.15%), and World health Organization (53.12%) definitions.

**Conclusion:** Almost two out of three type 2 diabetic patients in Sub-Saharan African countries have metabolic syndrome, which implies that its prevalence is high in patients with T2DM. Therefore, Policymakers (FMoH) need to design efficient strategies and guideline to reduce and control the burden of metabolic syndrome and its impact among diabetic population.

## Background

Metabolic syndrome (MetS) is becoming a serious global public health problem [1]. The metabolic syndrome is a clustering of key cardiovascular risk factors and includes abdominal obesity, dyslipidemia, hyperglycemia and hypertension with an individual[2, 3]. It is a complex disorder represented by a set of cardiovascular risk factors that are commonly associated with central adiposity and insulin resistance [4]. The magnitude of metabolic syndrome in developing countries is rising due to the global epidemic of obesity and type 2 diabetes mellitus [5, 6]. One study showed that the prevalence of metabolic syndrome was 18% among adult population in Sub-Saharan Africa [7].

The etiology of metabolic syndrome is complex and rooted in multiple risk factors. Studies suggest that numerous risk factors are responsible for metabolic syndrome in T2DM patients including old age[8–10], obesity [8, 9, 11], waist circumference[8, 12], systolic blood pressure[8, 10, 12], diastolic blood pressure [8, 10], triglyceride [8, 10, 12], low high density lipoprotein[8, 12, 13], family history of diabetes [9, 11], female [9, 10, 12], high low density lipoprotein [10], fasting blood sugar [12], upper socioeconomic status and urban[11], and HbA1c values and microalbuminuria[14].

Metabolic syndrome is a group of interconnected clinical and biological abnormalities which leads to cardiovascular diseases (CVDs)[5, 15], and it is frequently occurs among people with T2DM [2, 16]. It is an independent clinical indicator of macrovascular and microvascular complications in diabetic patients including coronary heart disease, stroke and other diseases involving the blood vessel. In addition it markedly increases health care cost, cardiovascular mortality, decreased quality of life, and increases the risk of premature death, renal disease, mental disorders and cancer [8, 13, 17–19].

The prevalence of metabolic syndrome in patients with T2DM varies widely across many literatures. For instance, the prevalence of metabolic syndrome is 57% in India [20], 70.4% in Iran [21], 84.8 in Malaysia [22], 77.9% in Korea [23], 51% Nigeria [24], 46.5% South Africa [25], 73% Zambia [26], 43% Zimbabwe [27], 68.6% Ghana [28], and 70.1% in Ethiopia [29]. Though different primary studies in Sub-Saharan African countries showed the prevalence of metabolic syndrome in patients with T2DM, their results have demonstrated substantial variation regarding its prevalence in the region. Therefore, this study was aimed to estimate the pooled prevalence of metabolic syndrome in patients with T2DM in Sub-Saharan African countries. The findings of this study would serve as a benchmark for policymakers to implement appropriate preventative measures, and to alleviate the double burden of metabolic syndrome.

## Methods

### Design and search strategy

Initially, Joanna Briggs Institute, and PROSPERO databases were searched to check out whether systematic review and meta-analysis is exist or for the presence of ongoing projects related to the current topic. The literatures were searched using PubMed, Scopus, Google scholar, African Journals Online, Web of Science, and Wiley Online Library. Manual search was done for grey literature available on local university shelves and institutional repositories. Moreover, the reference lists of all retrieved articles were conducted to identify additional relevant research to minimize publication bias to possible level. This search involved articles published from inception to April 27, 2020. The search was restricted to full texts, free articles, human studies, and English language publications. Endnote X 8.1 reference manager software was used to search, collect, organize search outcomes and for removal of duplicate articles. The search was conducted using the following medical subject headings (MeSH), and free-text terms: “metabolic syndrome”, “syndrome X”, “insulin resistance syndrome”, “prevalence”, “Type 2 diabetes mellitus”, “type 2 diabetes”, “diabetes Mellitus”, “non-insulin dependent diabetes”, “adult onset diabetes”, “Sub Saharan Africa”, and “names of each Sub Saharan Africa countries”. Boolean operators like “AND” and “OR” were used to combine search terms (Table 1).

**Table 1.** Pubmed search history.

| Search | Search terms | Hits |
| --- | --- | --- |
| 1 | Type 2 diabetes mellitus OR type 2 diabetes OR diabetes Mellitus OR non-insulin dependent diabetes OR adult onset diabetes | 92,317 |
| 2 | metabolic syndrome OR syndrome X OR insulin resistance syndrome | 41,625 |
| 3 | #1 and #2 | 4,150 |
| 4 | African filter((((Angola OR Benin OR Botswana OR“Burkina Faso”OR Burundi OR Cameroon OR“Cape Verde” OR“Central African Republic” OR Chad OR Comoros OR Congo OR“Democratic Republic of Congo” OR Djibouti OR“Equatorial Guinea” OR Eritrea OR Ethiopia OR Gabon OR Gambia OR Ghana OR Guinea OR“Guinea Bissau” OR “Ivory Coast” OR“Cote d’Ivoire” OR Kenya OR Lesotho OR Liberia OR Madagascar OR Malawi OR Mali OR Mauritania OR Mauritius OR Mozambique OR Namibia OR Niger OR Nigeria OR Principe OR Reunion OR Rwanda OR“Sao Tome” OR Senegal OR Seychelles OR“Sierra Leone” OR Somalia OR“South Africa” OR Sudan OR Swaziland OR Tanzania OR Togo OR Uganda OR“Western Sahara” OR Zambia OR Zimbabwe OR“Central Africa” OR“Central African” OR“West Africa” OR“West African” OR“Western Africa” OR“Western African” OR“EastAfrica” OR“East African” OR“Eastern Africa” OR “Eastern African” OR“South African” OR“Southern Africa” OR “Southern African” OR“sub Saharan Africa” OR“sub Saharan African” OR“sub Saharan Africa” OR“sub Saharan African” NOT“guinea pig” NOT“guinea pigs” NOT“aspergillus niger” )))) | 83,769 |
| 5 | #3 and #4 | 81 |
| 6 | Limits: from inception to 4/27/2020, studies done in Humans and English language | 81 |

### Eligibility criteria

Studies were included if they fulfilled the following criteria: (1) All observational studies which report the prevalence of metabolic syndrome in patients with T2DM; (2) articles published in peer-reviewed journals or grey literature; and (3) articles published in English from inception to April 27, 2020. Furthermore, if different diagnostic criteria of MetS were found in a study, our first choice was the National Cholesterol Education Program Adult Treatment Panel (NCEPATP III), and our second choice considered International Diabetes Federation (IDF). Studies were excluded if: (1) they were not fully accessible; (2) they possessed a poor quality score as per the stated criteria; (3) case series, letters, comments and editorials; and/or (4) they failed to measure the desired outcome (i.e., metabolic syndrome).

### Outcome of interest

The main outcome of interest was the prevalence of metabolic syndrome in patients with T2DM reported in the original paper both as percentage and as the number of metabolic syndrome (n) / total number of patients T2DM (N). The diagnostic criteria of metabolic syndrome in the current review included; International Diabetes Federation (IDF) [30], World Health Organization (WHO) [31], and the National Cholesterol Education Program Adult Treatment Panel III (NCEP ATP II) [32].

### Data extraction and quality assessment

Two authors independently extracted all necessary data from each study using Microsoft Excel spread sheet. If discrepancies between data extractors were observed, a third author was involved. For each included study, the following data were extracted: corresponding author, publication year, country, study design, sample size, data collection period, sampling technique, metabolic syndrome diagnostic criteria, response rate, and prevalence of metabolic syndrome with its 95% confidence interval (CI). The methodological quality of each included study was assessed using the Newcastle-Ottawa scale (NOS)[33]. This scale has several key criteria including the representativeness of the sample, response rate, measurement tool used, comparability of the subject, and the appropriateness of the statistical test used to analyze the data. Studies were included in the analysis if they scored ≥5 out of 10 points in three domains of ten modified NOS components for observational studies [34]. Furthermore, quality assurance checks were independently performed by three authors. Any disagreements at the time of data abstraction were resolved by discussion and consensus (Supplementary file 1).

### Assessment of risk of bias in included studies

The risk of bias of selected articles were assessed using the risk of bias tool for prevalence studies developed by Hoy et al. [35]. After reviewing different literatures, we decide that articles scored 8 or more “yes” answers out of ten point scale were considered to be low risk of bias; 6 to 7 “yes” answers were considered as moderate risk of bias; 5 or fewer “yes” answers were considered to be high risk of bias (see supplementary file 2). Two authors carried out the risk of bias assessment of the included studies independently. A summary of the areas considered in the assessment of each domain is included in the risk of bias assessment of included studies.

### Heterogeneity and publication bias

Heterogeneity between study estimates was assessed using the Cochran’s Q and the I^2^ statistic [36]. The I^2^ statistic estimates the percentage of total variation across studies due to true between-study differences rather than chance. In this study, heterogeneity was interpreted as an I^2^ value of 0% = no heterogeneity, 25% = low, 50% = moderate, and 75% = high [37]. In case of high heterogeneity, subgroup analysis to reduce heterogeneity, and meta regression analysis to explore sources of heterogeneity were employed. In addition, potential outliers were investigated in sensitivity analysis by dropping each study at a time. Potential publication bias was assessed by visually inspecting funnel plots and objectively using Egger test p< 0.05 were considered as statistical significant publication bias [38]. Trim-and-fill analysis was used to adjust estimates for the effects of publication bias.

### Statistical analysis

To obtain the meta-weighted prevalence of metabolic syndrome among T2DM, a meta-analysis using random-effects model DerSimonian and Laird method was utilised due to significant heterogeneity among studies (I^2^ = 87.8%,p<0.001)[39]. The pooled effect size (i.e. Prevalence) with a 95% confidence interval (CI) was generated and presented using a forest plot. For pooled data analysis, we merged the estimates of metabolic syndrome by NCEP ATP II, IDF, and WHO definitions criteria (average prevalence across those with multiple definitions). All data manipulation and statistical analyses was performed using the STATA™ Version 14 software [40].

### Presentation and reporting of results

The results of this review were reported based on the Preferred Reporting Items for Systematic Review and Meta-Analysis statement (PRISMA) guideline[41]. The entire process of study screening, selection, and inclusion were described with the aid of a flow diagram. Results were presented using forest plots and summary tables.

## Results

### Search results

Our compressive search strategy owns us a total of 1,325 articles. Of these, 1,318 articles were retrieved from PubMed (81), Scopus (299), Google Scholar (442), Web of Science (96), Wiley Online Library (318), and African Journals Online (159). The remaining 7 were found through a manual search. After excluding duplicate publications, 687 articles remained. About 565 articles were excluded after reading the titles and abstracts based on the pre-defined eligibility criteria. Out of them 122 articles were included and screened for further assessment. Finally, 23 articles included in the analysis (Fig. 1).

**Figure 1.**
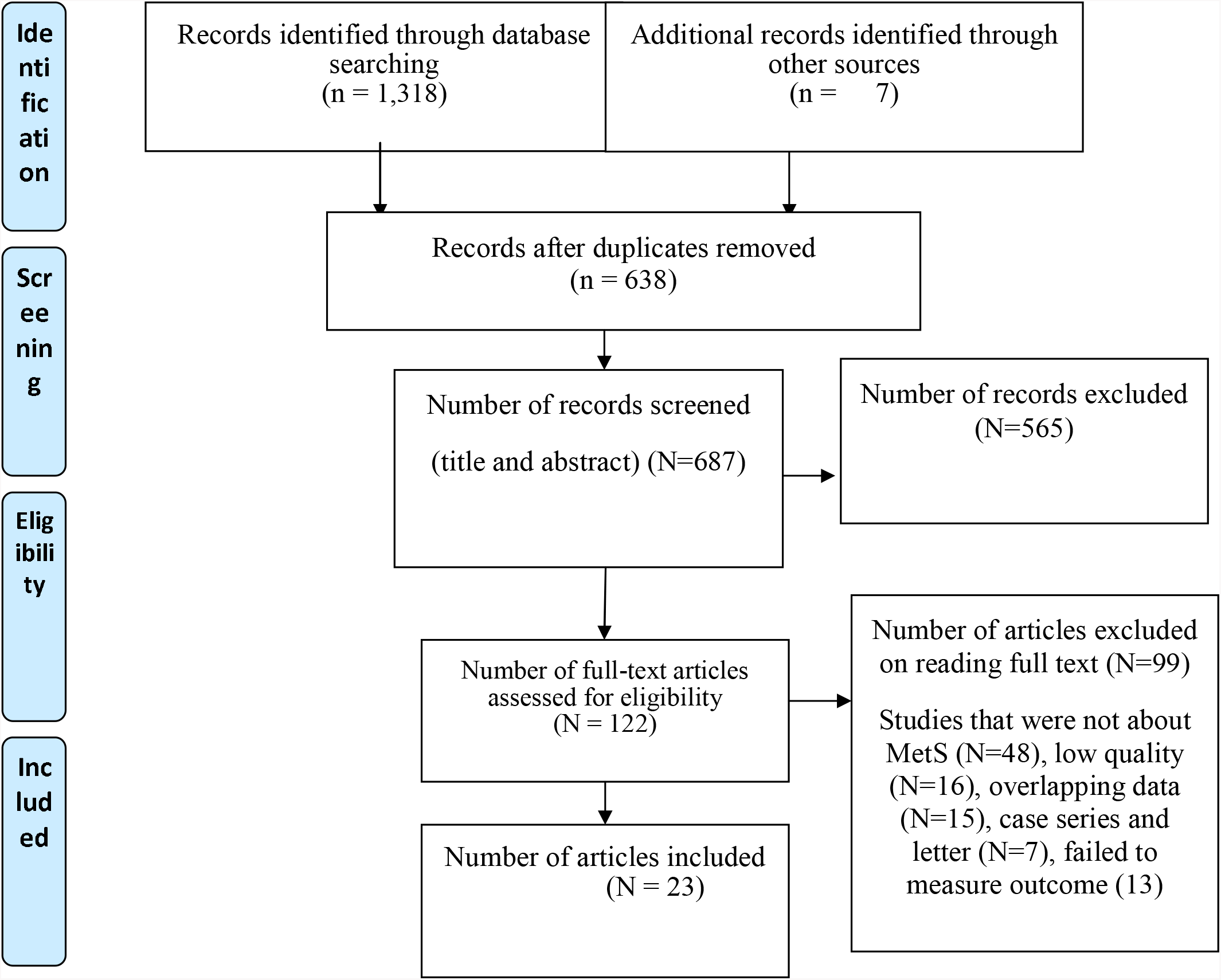
PRISMA flow chart of the study selection

### Baseline characteristics of the included studies

The search strategy yields a total of 23 studies for data extraction with a total sample of 6,482 T2DM patients which were found in sub-Saharan Africa. From the studies which included for the final analysis, the majority (95%) of the studies included were cross-sectional in study design. The total number of participants per study ranged from 103 to 634. The highest (90.6%), and the lowest (24%) prevalence of metabolic syndrome were reported in the studies conducted in Ghana [42, 43]. Four studies were conducted in Ethiopia [29, 44–46], eight in Ghana [42, 43, 47–52], seven in Nigeria [11, 24, 53–57], one each from Zambia [26], South Africa [25], Zimbabwe [27], and Cameroon[58]. Regarding the diagnostic criteria used to assess metabolic syndrome in patients with T2DM, five studies [11, 24, 27, 53, 54] used the WHO definition, eight studies [25, 43, 44, 46, 47, 49, 56, 58] used IDF, and ten studies [26, 29, 42, 45, 48, 50–52, 55] were used NCEP-ATP III. The quality score of each primary study, based on the Newcastle-Ottawa quality score assessment was moderate to high for all 23 articles assessed.

**Table 2.**
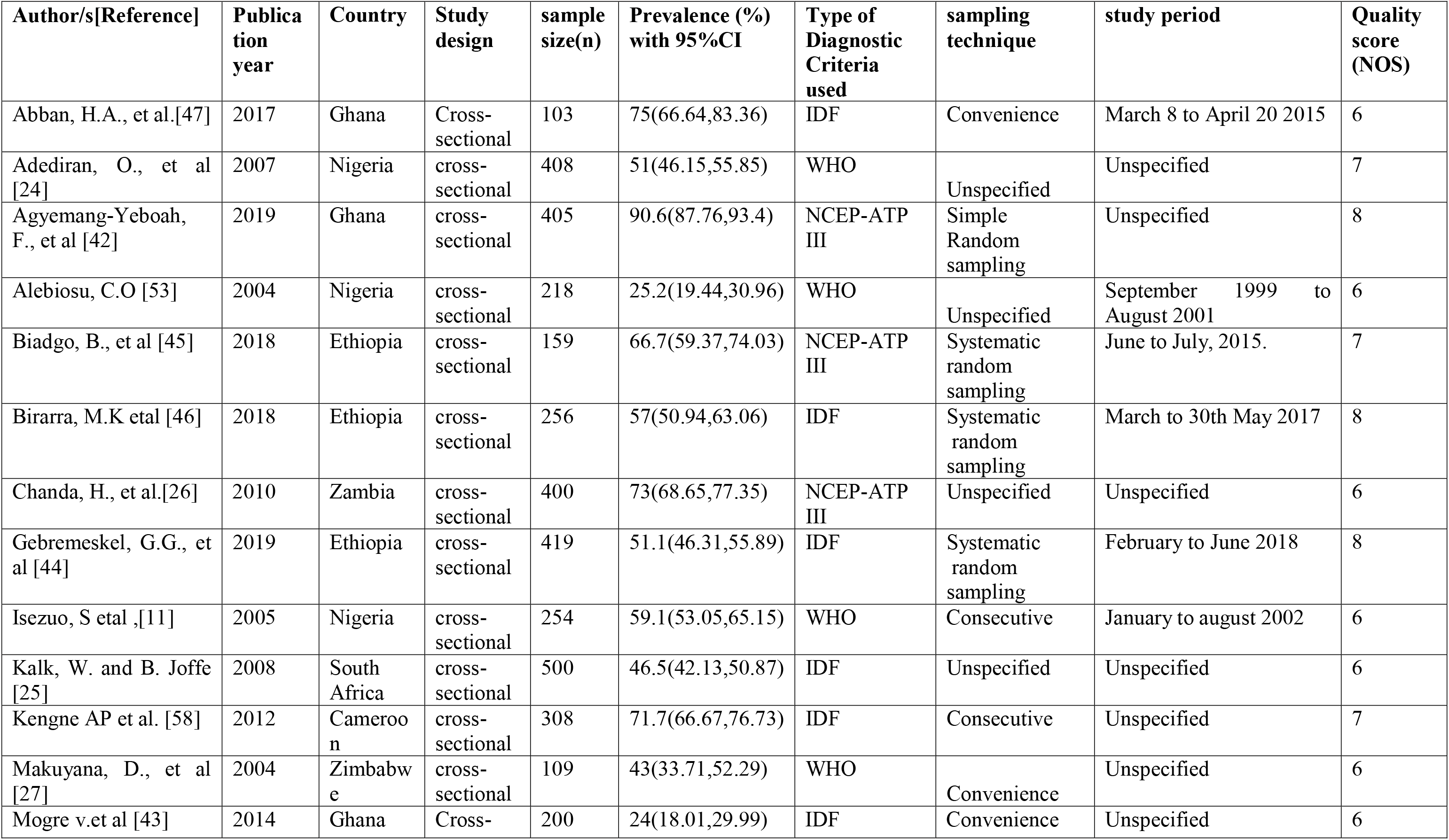
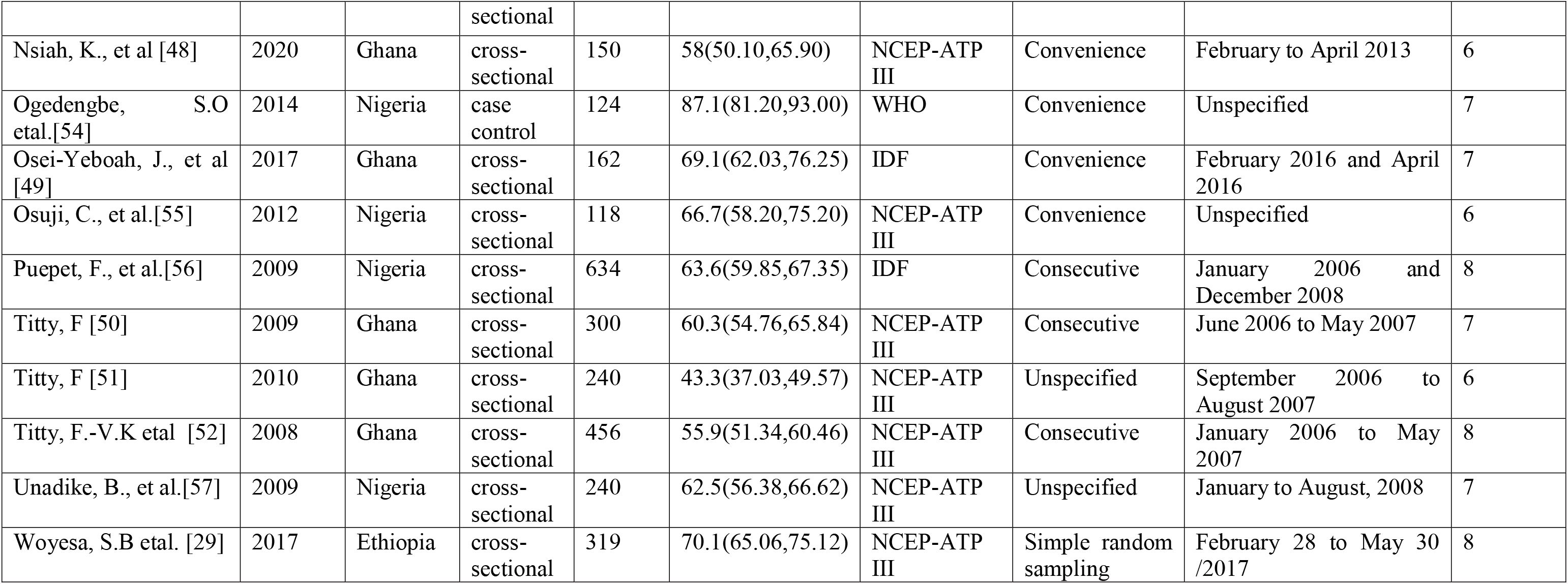
Baseline characteristics of the included studies.

### Prevalence of metabolic syndrome

The current meta-analysis using the random effects model showed that the pooled prevalence of metabolic syndrome in patients with T2DM was 59.62% (95% CI: 52.20, 67.03) with substantial level of heterogeneity (I^2^ = 87.8%; p <0.001), (Fig. 2).

**Figure 2.**
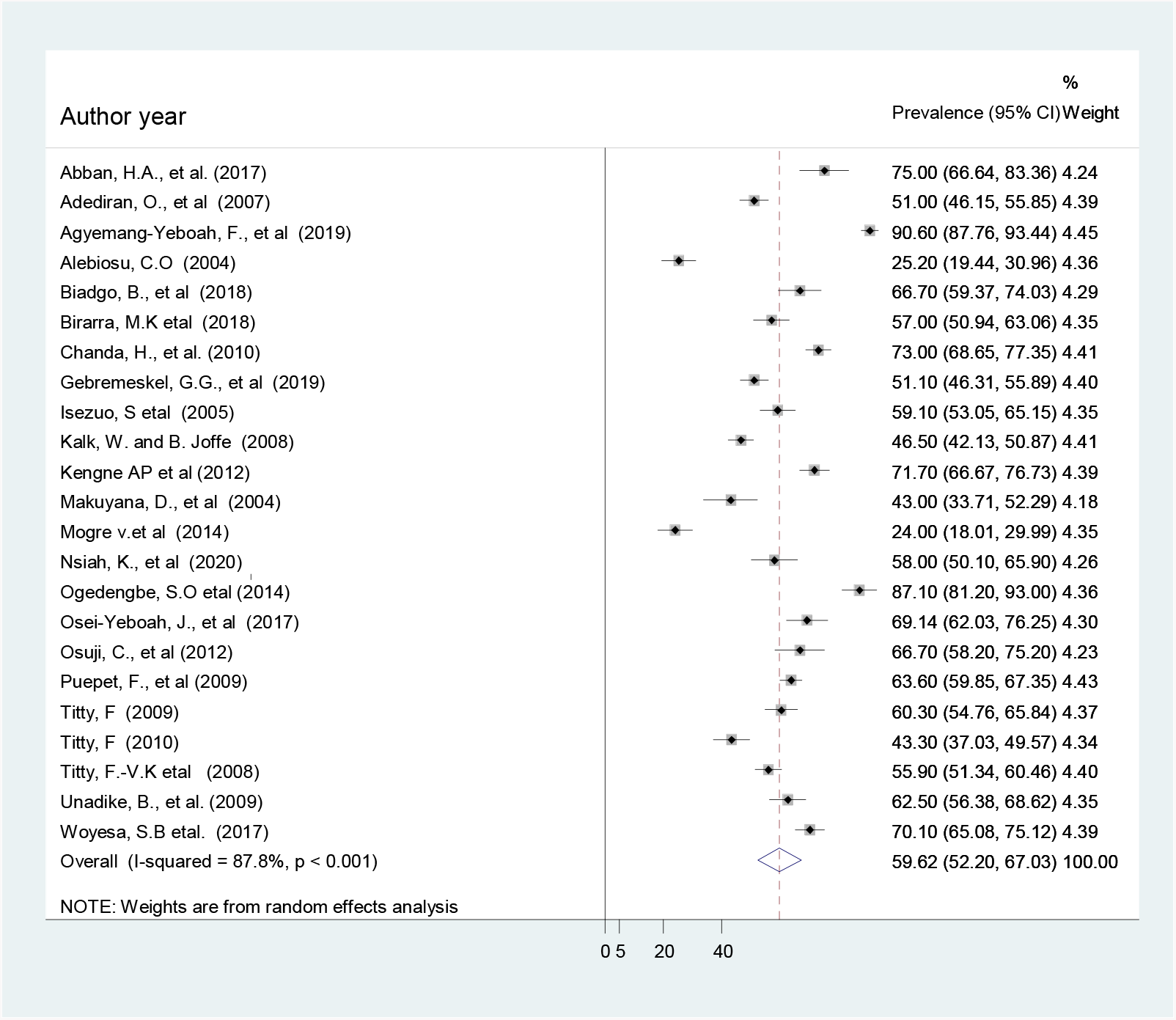
Pooled prevalence of metabolic syndrome in patents with T2DM

**Figure 3.**
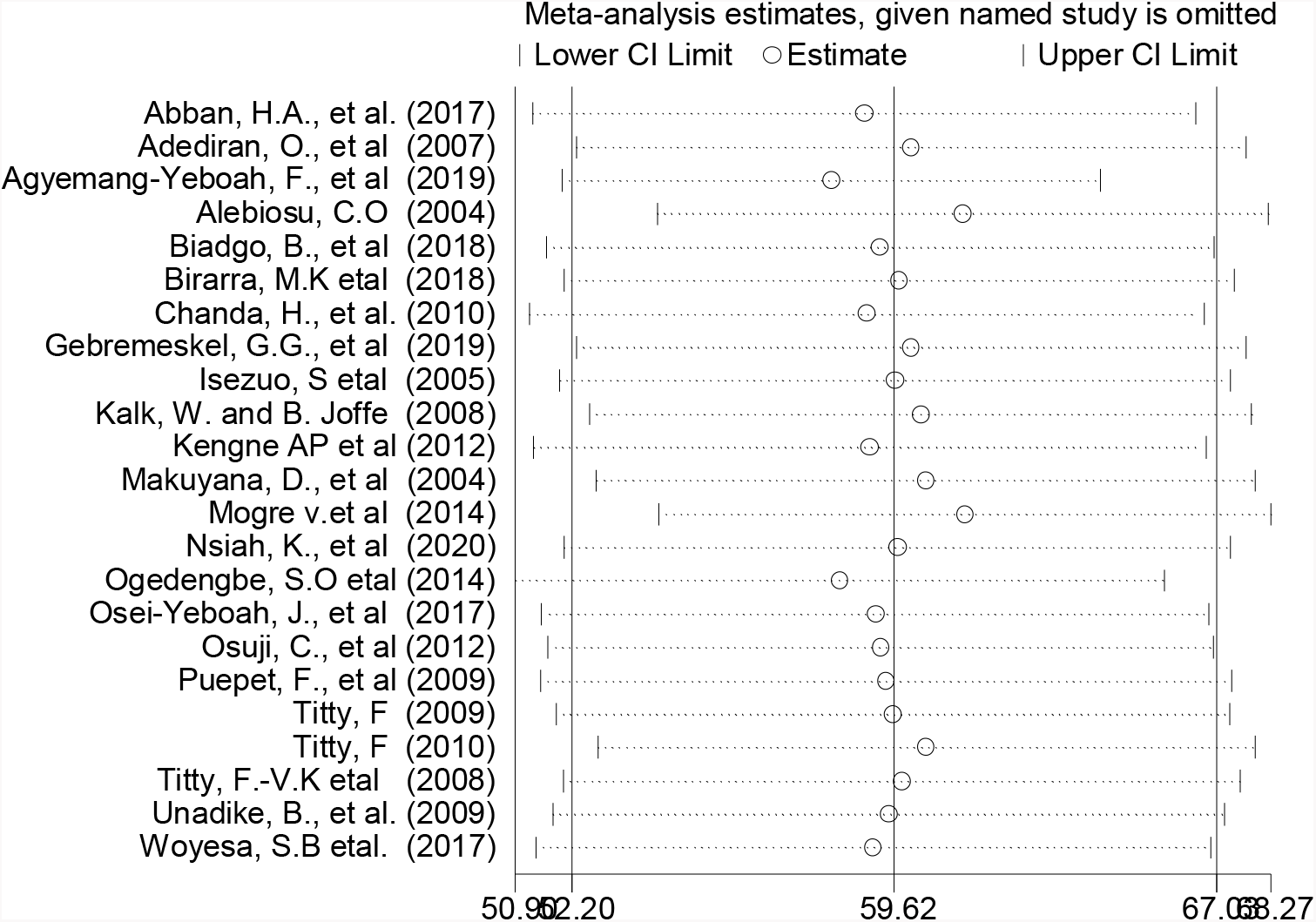
Result of sensitivity analysis of the 23 studies.

### Subgroup analysis

To identify the source of heterogeneity across the included studies, subgroup analysis was deployed using publication year, country, diagnostic criteria, and sampling technique Based on the subgroup analysis result, the pooled prevalence of metabolic syndrome was 65.62% in studies published after 2010, 61.14% in Ethiopian studies, 64.8% among studies using NCEP ATP II as a diagnostic criteria, 80.46% in studies with simple random sampling technique (Table 2).

**Table 2.** The results of subgroup analysis by different category of the studies.

| Sub-group | Category | No. of studies | Sample size | Prevalence (95% CI) | p-value | I<sup>2</sup> |
| --- | --- | --- | --- | --- | --- | --- |
| *By year of publication* | ≤ 2010 | 11 | 3,759 | 53.17(45.57, 60.77) | <0.001 | 88% |
|  | > 2010 | 12 | 2,723 | 65.62(54.23,77.01) | <0.001 | 85.5% |
| *Country* | Ethiopia | 4 | 1,153 | 61.14(51.74,70.53) | <0.001 | 82.9% |
|  | Ghana | 8 | 2,016 | 59.54(42.53,76.56) | <0.001 | 88.7% |
|  | Nigeria | 7 | 1,996 | 59.27(46.08,72.47) | <0.001 | 87.5% |
|  | Others | 4 | 1,317 | 58.80(43.42,74.18) | <0.001 | 87.1% |
| *Diagnostic criteria* | IDF | 8 | 2,582 | 57.15(46.69,67.61) | <0.001 | 86.8% |
|  | WHO | 5 | 1,113 | 53.12(32.69,73.55) | <0.001 | 88.2% |
|  | NCEP ATP II | 10 | 2,787 | 64.80(54.74,74.86) | <0.001 | 87.3% |
| *Sampling technique* | Systematic random sampling | 3 | 834 | 57.91(49.32,66.49) | 0.008 | 73.8% |
|  | Simple random sampling | 2 | 724 | 80.46(60.37,100.55) | <0.001 | 87.9% |
|  | Consecutive | 5 | 1,952 | 62.16(59.93,67.40) | 0.07 | 72.6% |
|  | Convenience | 7 | 966 | 60.43(42.34,78.41) | <0.001 | 87.6% |
|  | Not specified | 6 | 2,006 | 50.29(37.05,63.53) | <0.001 | 87.5% |

Others; Zambia, South Africa, Zimbabwe, and Cameroon

**Table 3.**
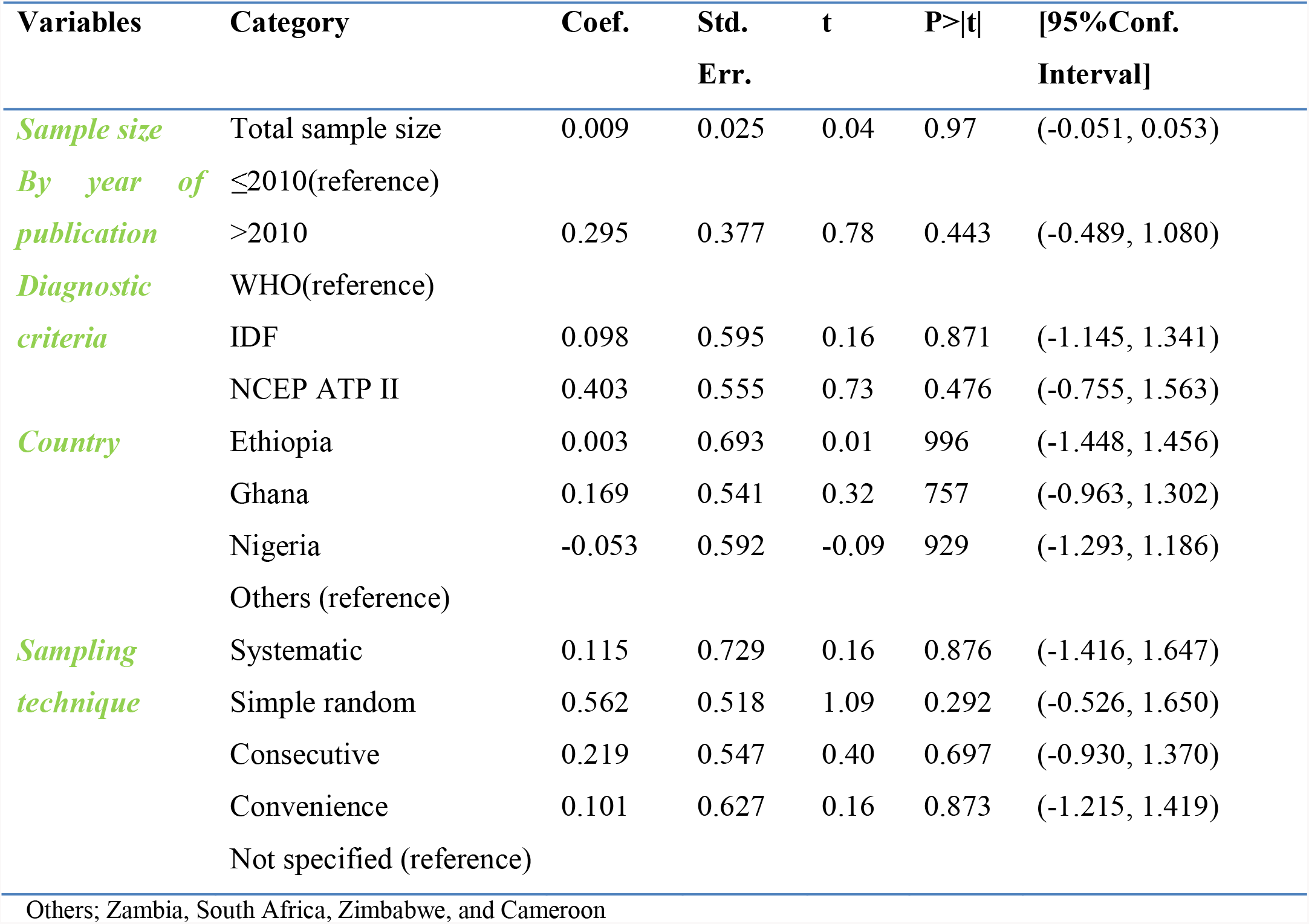
Meta-regression analysis for the included studies to identify the source(s) of heterogeneity.

| Variables | Category | Coef. | Std.<br>Err. | t | P>\|t\| | [95%Conf.<br>Interval] |
| --- | --- | --- | --- | --- | --- | --- |
| *Sample size* | Total sample size | 0.009 | 0.025 | 0.04 | 0.97 | (-0.051, 0.053) |
| *By year of publication* | ≤2010(reference) |  |  |  |  |  |
|  | >2010 | 0.295 | 0.377 | 0.78 | 0.443 | (-0.489, 1.080) |
| *Diagnostic criteria* | WHO(reference) |  |  |  |  |  |
|  | IDF | 0.098 | 0.595 | 0.16 | 0.871 | (-1.145, 1.341) |
|  | NCEP ATP II | 0.403 | 0.555 | 0.73 | 0.476 | (-0.755, 1.563) |
| *Country* | Ethiopia | 0.003 | 0.693 | 0.01 | 996 | (-1.448, 1.456) |
|  | Ghana | 0.169 | 0.541 | 0.32 | 757 | (-0.963, 1.302) |
|  | Nigeria | -0.053 | 0.592 | -0.09 | 929 | (-1.293, 1.186) |
|  | Others (reference) |  |  |  |  |  |
| *Sampling technique* | Systematic | 0.115 | 0.729 | 0.16 | 0.876 | (-1.416, 1.647) |
|  | Simple random | 0.562 | 0.518 | 1.09 | 0.292 | (-0.526, 1.650) |
|  | Consecutive | 0.219 | 0.547 | 0.40 | 0.697 | (-0.930, 1.370) |
|  | Convenience | 0.101 | 0.627 | 0.16 | 0.873 | (-1.215, 1.419) |
|  | Not specified (reference) |  |  |  |  |  |

Others; Zambia, South Africa, Zimbabwe, and Cameroon

### Meta regression analysis

To identify the source(s) of heterogeneity, meta-regression analysis was undertaken by considering year of publication, sample size, diagnostic criteria, country, and sampling technique. However, our results showed that all covariates were not statistically significant for the presence of heterogeneity (Table 4).

### Sensitivity analysis

In sensitivity analyses using the leave-one-out approach, excluding none of the studies had a significant effect on the pooled prevalence estimates and measures of heterogeneity within primary studies. Therefore, sensitivity analyses using the random effects model revealed that no single study influenced the overall prevalence of metabolic syndrome in patient with type 2 DM (Fig. 4).

**Figure 4.**
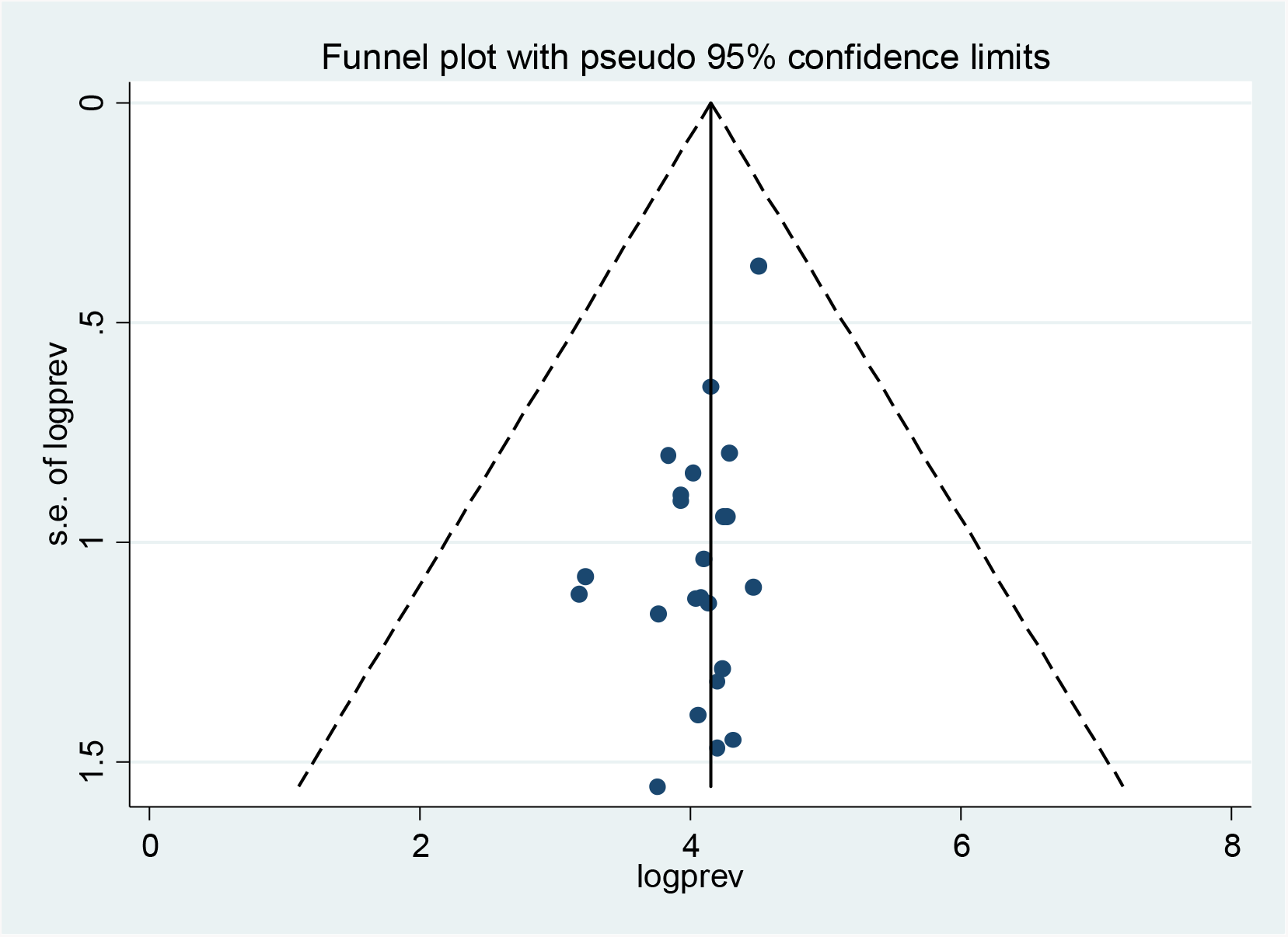
Funnel plot to test publication bias of the 23 studies.

### Publication bias

The visual inspection of the funnel plot showed that there was no publication bias among the included studies, as illustrated by the symmetrical distribution of the funnel plot (Fig. 4). However, the result of Egger test was statistically significant for the presence of publication bias (P = 0.006). Therefore, to reduce and adjust publication bias, trim and fill analysis was performed (Fig. 5). Trim and fill analysis is a nonparametric method for estimating the number of missing studies that might exist [59]. Though we have carried out the pooled prevalence of the MetS after trim and fill analysis, the finding was similar.

**Figure 5.**
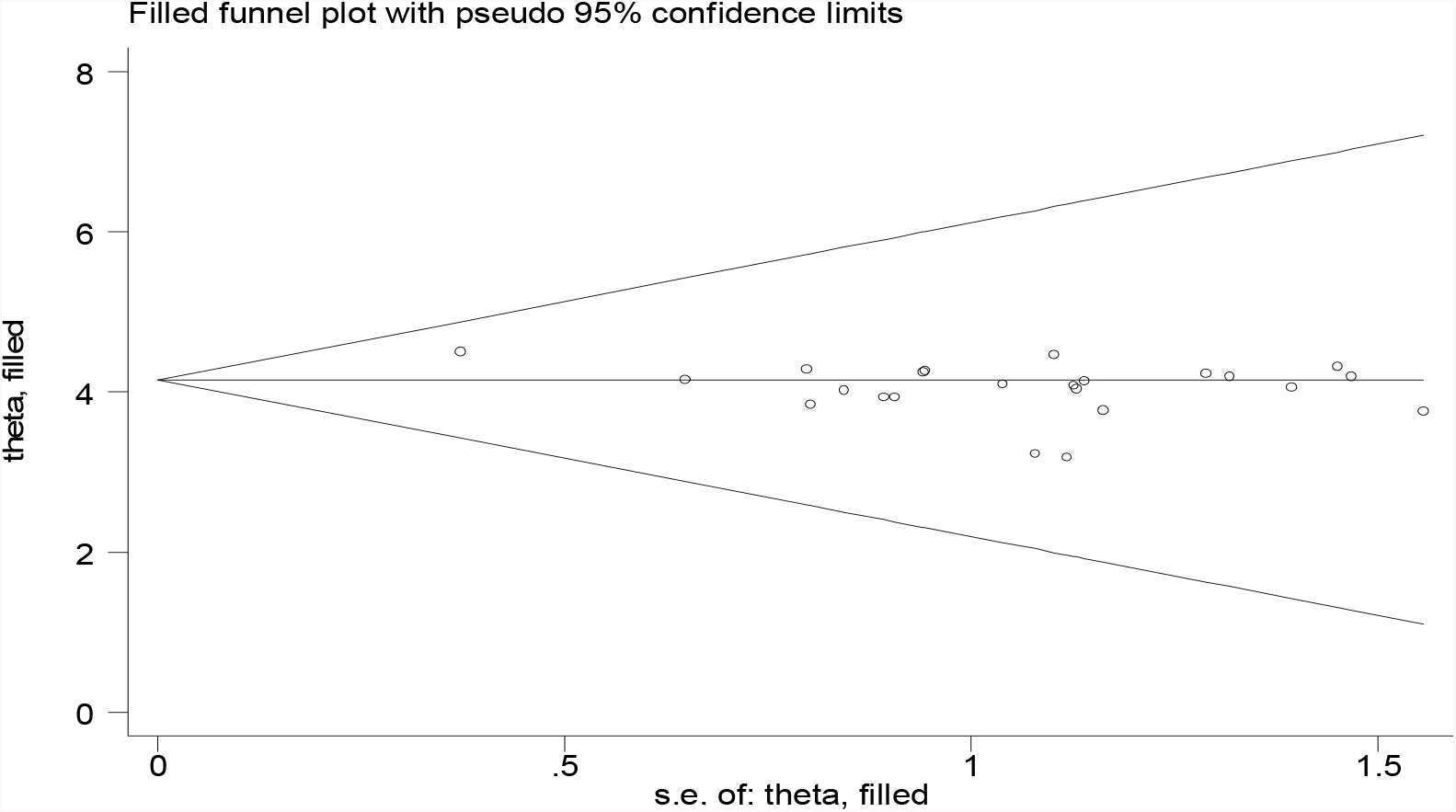
Result of trim and filled analysis for adjusting publication bias of the 23 studies.

## Discussion

In this meta-analysis the pooled prevalence of metabolic syndrome in patients with T2DM in Sub-Saharan Africa was estimated to be 59.62% (95% CI: 52.20, 67.03) irrespective of the diagnostic criteria. The overall estimated prevalence of metabolic syndrome among T2DM patients in Sub-Saharan Africa was ranged between 24% and 90.6%. This result was substantially higher than a systematic review conducted in different European countries where the prevalence ranged between 3% and 71.7% [60]. This variation could be justified by difference in diagnostic criteria, variation by ethnicity, possible cultural differences [43], health care seeking behavior’s between population, and differences in life style of study participants. In addition, this high prevalence of metabolic syndrome in the SSA could be due to epidemiological transition which has placed them on a double burden of disease, and socioeconomic factors like sedentary lifestyle and eating habits [10, 61].

Based on the subgroup analysis by country, the highest prevalence of metabolic syndrome among T2DM in Sub-Saharan African countries observed in Ethiopia (61.14% (95% CI: 51.74, 70.53)), followed by Ghana (59.54% (95% CI: 42.53, 76.56)). This variation might be due the difference in the availability of healthcare resources in the countries, and the diagnostic methods used. Based on the diagnostic criteria used (NCEP ATP II, IDF, WHO) across the included studies, the highest prevalence of metabolic syndrome (64.8 % (955CI: 54.74, 74.86)) was reported across studies using the NCEP ATP III diagnostic criteria and the lowest prevalence (53.12% (95%CI: 32.69, 73.55)) of metabolic syndrome was reported across studies using the WHO-definition as a diagnostic criteria. A similar findings in variation of metabolic syndrome prevalence per diagnostic criteria is also reported in a study conducted different European countries, where the prevalence of MetS using NCEP-ATP III criteria ranged from 2% to 56%, using IDF ranged from 19 % to 60 %, and only 2 % was found based on WHO-definition [60]. Another similar study done Sri Lankan revealed that the prevalence of MetS varied significantly between different diagnostic criteria. They found that the Prevalence of MetS was highest by WHO definition (70%) followed by IDF (44%) and NCEP-ATP III (29%) definitions[62]. These data suggest that while the prevalence of MetS differs between different diagnostic criteria in all populations, the extent of the differences may be larger in some populations than in others [63].

This study has implications for clinical practice. Determining the prevalence of metabolic syndrome among type 2 diabetic patients is critical to guide healthcare professionals to minimize the risk of metabolic syndrome by providing guidance to the patient who has undergone diabetic care follow up. Moreover, it gives information about the burden and public health impact of metabolic syndrome in the region for possible consideration during routine diabetic patient care.

The current meta-analysis has limitations that should be considered in the future research. First, studies from few countries in the sub-Saharan Africa included in this meta-analysis may be difficult to generalize the findings to all type 2 diabetic patients in the region. Second, different criteria used to diagnose metabolic syndrome in the included studies may affect the estimation of the pooled prevalence. Third, this study does not identify the predictors of metabolic syndrome among patients with T2DM.

## Conclusion

Almost two out of three of type 2 diabetic patients in Sub-Saharan African countries have metabolic syndrome, which implies that it is highly prevalent in patients with T2DM. Its prevalence varies across countries in the region with the highest prevalence in Ethiopia. Therefore, situation-based interventions and country context-specific preventive strategies should be developed to reduce the burden of metabolic syndrome. Further research is needed to identify possible risk factors associated with MetS in patients with T2DM in SSA populations to assist the prevention efforts.

## Data Availability

All relevant data are within the paper and it’s supporting information files. There is no separate data set to share

## List of abbreviations

IDF: International diabetic federation
Mets: metabolic syndrome
NCEP ATP II: National Cholesterol Education Program Adult Treatment Panel III
T2DM: type 2 diabetes mellitus
WHO: world health organization

## Declaration

### Ethics approval and consent to participate

Not applicable.

### Consent for publication

Not applicable.

### Authors’ contributions

WSS, YAA, WM, and TYA developed the protocol and were involved in the design, selection of study, data extraction, statistical analysis, and developing the initial drafts of the manuscript.

HM, GD, AM, DA, and MG were involved in data extraction, quality assessment, statistical analysis and revising. WSS, YAA, GD, DA, and HM prepared and edited the final draft of the manuscript. All authors read and approved the final draft of the manuscript.

## Acknowledgements

Not applicable

